# Characterizing the disproportionate burden of SARS-CoV-2 variants of concern among essential workers in the Greater Toronto Area, Canada

**DOI:** 10.1101/2021.03.22.21254127

**Authors:** Zain Chagla, Huiting Ma, Beate Sander, Stefan D. Baral, Sharmistha Mishra

**Author notes:** equal contribution (Stefan D. Baral, Sharmistha Mishra).

## Abstract

**Importance:** The emergence of SARS-CoV-2 Variants of Concern (VOC) across North America has been associated with concerns of increased COVID-19 transmission. Characterizing the distribution of VOCs can inform development of policies and programs to address the prevention needs of disproportionately affected communities.

**Objective:** We compared per-capita rates of COVID-19 cases (overall and VOC) from February 3, 2021 to March 10, 2021, across neighborhoods in the health regions of Toronto and Peel, Ontario, by proportion of the population working in essential services and income.

**Design:** Descriptive epidemiological analysis, integrating COVID-19 surveillance and census data. Per-capita daily epidemic curves were generated using 7-days rolling averages for cases and deaths. Cumulative per-capita rates were determined using census-reported population of each neighbourhood.

**Setting:** The study setting was the city of Toronto and the region of Peel (the City of Brampton, Mississauga, and Caledon), Canada’s largest cities with a combined population of 4.3 million. This area of Canada has had one of the highest incident rates of COVID-19 throughout the pandemic.

**Participants:** We used person-level data on laboratory-confirmed COVID-19 community cases (N=22,478) and census data for neighborhood-level attributes.

**Exposures:** We stratified neighbourhoods, i.e., dissemination areas which represent geographic areas of approximately 400-700 individuals, into tertiles by ranking the proportion of population in each neighbourhood working in essential services (health, trades, transport, equipment, manufacturing, utilities, sales, services, agriculture); and the per-person equivalent household income.

**Main Outcome(s) and Measure(s):** The primary outcomes were laboratory-confirmed COVID-19 cases overall and VOC positives by neighbourhood.

**Results:** During the study period, VOC cases emerged faster in groups with lowest income (growth rate 43.8%, 34.6% and 21.6% by income tertile from lowest to highest), and most essential work (growth rate 18.4%, 30.8% and 50.8% by tertile from lowest tertile of essential workers to highest tertile of essential workers).

**Conclusions and Relevance:** The recent introduction of VOC in a large urban area has disproportionately affected neighbourhoods with the most essential workers and lowest income levels. Notably, this is consistent with the increased burden of non-VOC COVID-19 cases suggesting shared risk factors. To date, restrictive public health strategies have been of limited impact in these communities suggesting the need for complementary and well-specified supportive strategies to address disparities and overall incidence of both VOC and non-VOC COVID-19.

## Introduction

Variants of concern (VOC) of SARS-CoV-2 emerged towards the end of 2020, presenting new challenges on epidemic containment (1). Several VOCs have been identified, including VOC 202012/01 (B.1.1.7). The emergence of B.1.1.7 resulted in documented replacement of wild type strains in the United Kingdom and Denmark and was associated with an increase in secondary attack rates (1) and potential increase in severity (2). Administrative municipal, regional, or national bodies have responded to increased COVID-19-related morbidity and mortality, particularly with the emergence of VOC, with non-pharmaceutical interventions (NPI), such as masking, physical distancing, and closure of non-essential businesses, indoor dining, and schools.

Across countries, the SARS-CoV-2 epidemic prior to the emergence of VOC was disproportionately concentrated in neighbourhoods of low socioeconomic status, with higher burdens in lower income neighbourhoods and among front-line essential workers (3, 4). Higher density networks may be particularly at risk of VOC growth given the higher secondary attack rates associated with VOC (5). Thus, with the rapid emergence of VOC in Canada, we seek to compare the pattern and trajectory of VOC among lower-income and essential worker communities in the context of existing NPIs in the two of the country’s largest cities.

## Methods

We used Case and Contact Management Solutions (CCM)+ person-level data on VOC screen positive cases among laboratory-confirmed COVID-19 cases between February 3rd and March 10, 2021, and the Statistics Canada 2016 Census data for neighbourhood-level characteristics. Two public health units were included in the analysis, the City of Toronto (population 2.9 million), and the Region of Peel (population 1.4 million), the two most populous areas of Ontario, with the highest level of ethnic diversity and per capita cases of COVID-19 (6). During the time period included in this analysis, both regions had public health measures in place, including closed non essential businesses, restaurants, fitness centers, and limits to only interact with one’s household (7). From February 3 onwards, all samples positive on RT-PCR with a cycle threshold < 35 underwent Polymerase Chain Reaction (PCR) for the N501Y mutation as screening, and genetic sequencing for positive results (8). Neighourhoods were stratified into tertiles ranked by proportion of the population engaged in essential services (manufacturing, utilities, trades, transport, equipment, agriculture, sales, services, and health); and per-person equivalent income after tax. Tertile 1, 2, and 3 each represent approximately 33% of the total population; and comprise neighbourhoods where a median of 30.4% (inter-quartile range [IQR] 25.0-35.3%), 47.9% (IQR42.1%-52.0%), and 63.2% (IQR 59.4%-68.1%) of the population, respectively, worked in essential services; and a median income of $33,819 (29,337-37,054), 45,487 (41,411-48,079), and 60,652 (55,001-69,982) CAD. We compared the daily per-capita rates (7-day rolling average) of new cases and VOC screen positive cases, as well as the proportion of cumulative cases, by neighbourhood tertile. We excluded cases among residents of long-term care homes because they represent a different outbreak setting from community-level outbreaks and transmissions. Analyses were conducted in R (version 4.0.2).

## Results

A total of 19,912 cases were observed during the study period, of which 12,860 (64.6%) were screened for a VOC, and 5,084 (25.5%) screened positive. At the time of this reporting 93% of all samples which screened positive on N501Y PCR and sequenced, were the B.1.1.7 lineage (6). As shown in Figure 1A, per-capita cases are increasing in tertiles 2 and 3, with a smaller relative increase in tertile 1, representing the beginning of the region’s “third wave” in neighbourhoods with the highest concentration of essential workers.The growth rate of VOCs was 11.3%, 19.8%, and 30.8% in tertiles 1, 2, and 3 respectively, in the last week of the study period (Figure 1A). Between February 3 and March 10, 19.0%, 32.7%, and 48.3% of overall cases were in tertiles 1, 2, and 3 (Figure 1C); while 18.4%, 30.8% and 50.8% of VOC cases were tertiles 1, 2, and 3, respectively (Figure 1D).

**Figure 1.**
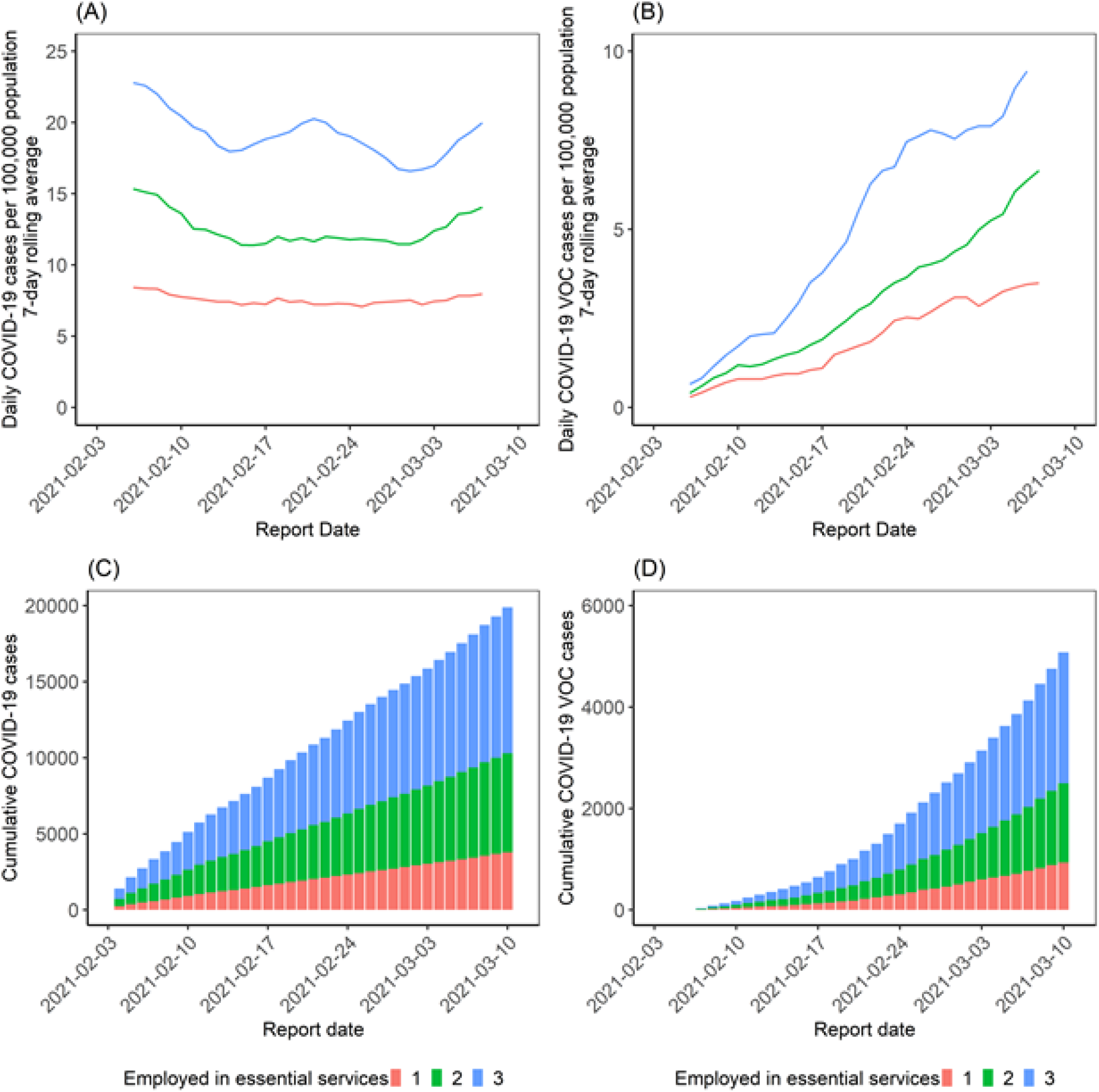
**Daily rates and cumulative COVID-19 overall and VOC cases in Toronto and Peel regions, Canada (February 3 - March 10, 2021), by neighbourhood-level prevalence of essential workers**. By March 7, the 7-day rolling average had reached 8.0, 14.1, and 20.0 per 100,000 population in tertiles 1,2, and 3 respectively (A); and VOC daily rates were at 3.5, 6.6, and 10.1 per 100,000 population in tertiles 1,2, and 3 respectively (B). Cumulative number of cases, stratified by tertiles are shown for overall cases (C) and VOC cases (D). VOC (variants of concern, detected via N501Y screening). Cases exclude those diagnosed among residents of long-term care homes.

A similar pattern was observed with income tertiles, with the third wave concentrated in lower income neighbourhoods (Figure 2A); and VOC growth rates of 26.2%, 21.1%, and 14.6% in tertiles 1, 2, and 3 respectively (Figure 1B). Within the study period, 43.8%, 35.6%, and 20.6% of overall cases were in tertiles 1, 2, and 3 (Figure 2C); while 43.8%, 34.6% and 21.6% of VOC cases were tertiles 1, 2, and 3, respectively (Figure 2D).

**Figure 2.**
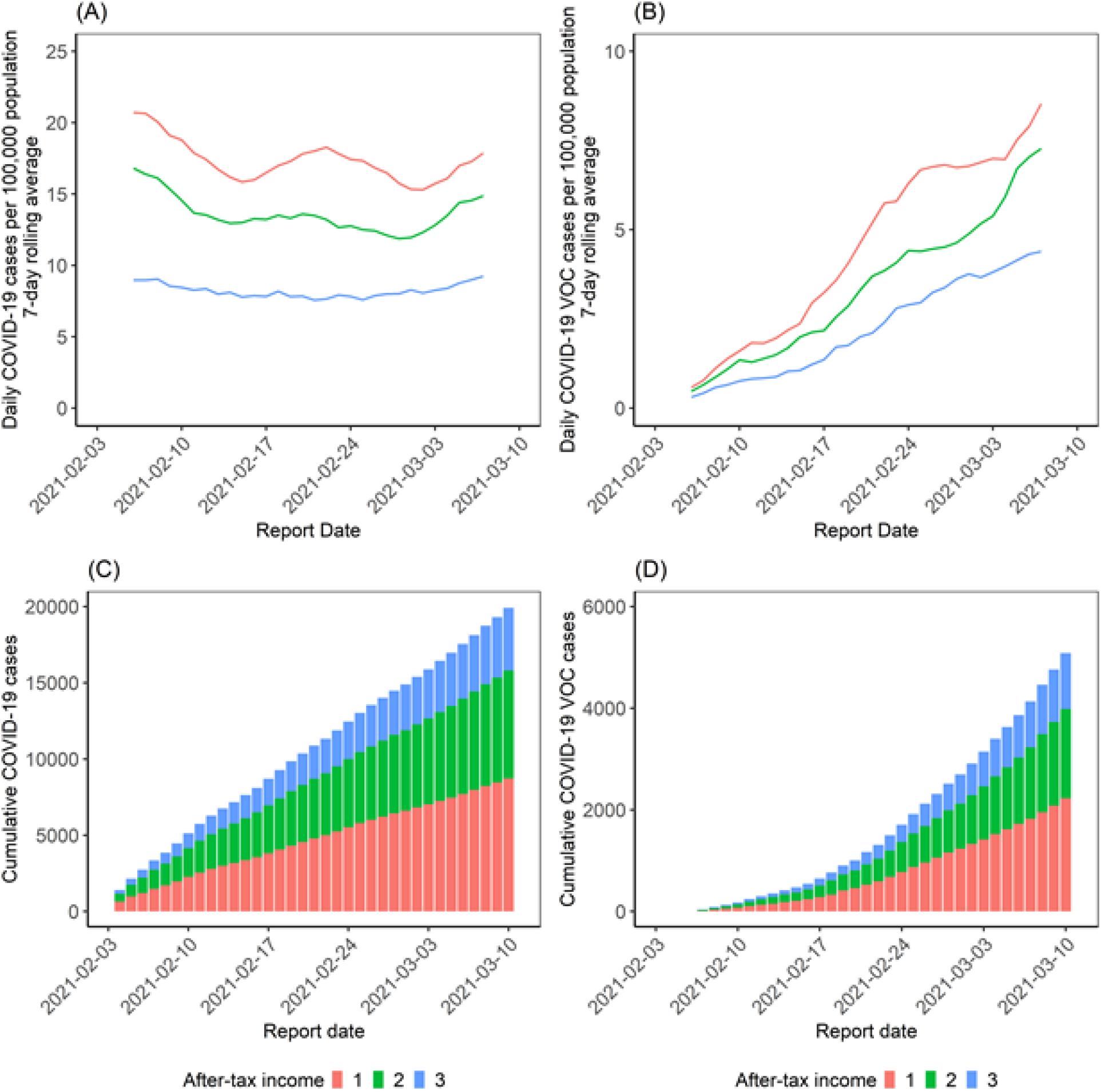
**Daily rates and cumulative COVID-19 overall and VOC cases in Toronto and Peel regions, Canada (February 3 - March 10, 2021), by neighbourhood-level household income tertile**. By March 7, the 7-day rolling average had reached 17.9, 14.9 and 9.2 per 100,000 population in tertiles 1,2, and 3 respectively (A); and VOC daily rates were at 8.5, 7.3, and 4.4 per 100,000 population in tertiles 1,2, and 3 respectively (B). Cumulative number of cases, stratified by tertiles are shown for overall cases (C) and VOC cases (D). VOC (variants of concern, detected via N501Y screening). Cases exclude those diagnosed among residents of long-term care homes. After-tax income tertile refers to neighbourhoods ranked by after tax, per-person equivalent household income.

## Discussion

Following the emergence and initiation of routine genomic surveillance for VOC in Canada, these results demonstrate that the epidemiology of the VOCs is similar to that of wild type SARS-CoV-2 in disproportionately affecting essential workers and in economically marginalized neighbourhoods (3,4). The recent VOC growth rate is faster in these neighbourhoods which may now be entering the “third wave” of the pandemic, showing a disproportionate burden of illness. The rapid emergence and concentration of VOCs has been described as a separate epidemic from that of non-VOC SARS-CoV-2. However in 2021, VOCs have rapidly concentrated in the same networks where both cases and associated morbidity and mortality have been concentrated throughout almost the entire COVID-19 pandemic suggesting prevention needs in these communities remain unmet. This is further supported by the fact that during the entirety of the study period, the Toronto and Peel region have been under a state of significant societal restrictions, with closures of non essential businesses, restaurants, bars, fitness centers, and legislation against public and private gatherings.

The finding that VOC are rapidly spreading through the same transmission networks as wild type SARS-CoV-2, and during a period of ongoing societal restrictions on mobility and shelter-in-place recommendations, suggests the need for targeted and tailored interventions to interrupt transmission chains in essential worker communities, including workplaces and households.

These include resources to improve quarantine and isolation including financial supports, paid leave, and voluntary temporary housing support. Moreover, interventions in essential businesses could include technical support for infection prevention and control in essential workplaces and outreach-based testing strategies adaptive to lived realities of essential workers. Finally, given the concentration of transmissions in the context of workplaces and their overlap with household density (3), vaccination at workplace and among subgroups with high rates of contact (i.e. essential workers) could be a prioritization strategy to mitigate transmissions and VOC growth (and thus, directly and indirectly reduce mortality) while also vaccinating those most at risk of severe outcomes if infected (9, 10)

The limitations of these analyses include the use of ecological data, rather than individual-level data on income and occupation for this analysis; and the potential for differential SARS-CoV-2 testing across tertiles, with proportionally less testing in lower-income neighbourhoods as shown previously (3) and thus, the potential for underestimating both wild-type in VOC cases in these communities.

Restrictive intervention strategies have represented the foundation of most COVID-19 public health interventions. Given that VOCs rapidly spread in the same transmission networks as wild-type SARS-CoV-2, and their emergence during a period of intensive societal restrictions, suggests focused measures in this population are needed. Moving forward necessitates ensuring that public health measures avoid reinforcing underlying inequities by leveraging resource and service-based strategies to minimize the future spread of novel VOC and associated morbidity and mortality.

## Data Availability

We used Case and Contact Management Solutions (CCM)+ person-level data on VOC screen positive cases among laboratory-confirmed COVID-19 cases between February 3rd and March 10, 2021. Such data are not currently publicly available. The analysis code, aggregated output data, and figures of daily per-capita and cumulative epidemic curves are publicly available on GitHub: https://git.io/J3m6r. We used Statistics Canada 2016 Census data for neighbourhood-level characteristics and these data are publicly available.

## Ethics approval

The University of Toronto Health Sciences Research Ethics Board (protocol no. 39253) approved the study.

## Acknowledgements

Reported COVID-19 cases were obtained from the Contact Management Solutions (CCM)+ via the Ontario COVID-19 Modelling Consensus Table and Ontario Ministry of Health. We thank Kristy Yiu (MAP Centre for Urban Health Solutions, St. Michael’s Hospital) for support with data management, and the Ontario Community Health Profiles Partnership.

## Competing Interest Statement

The authors have declared no competing interest.

## Funding Statement

This work was funded by the Canadian Institutes of Health Research (grant no. VR5-172683).

## Notes

### Author Declarations

Ethics approval The University of Toronto Health Sciences Research Ethics Board (protocol no. 39253) approved the study.

### Summary of Updates

Updated Data Availability Statement and cleaned up formatting issues in manuscript

